# “Exponere” A National Level Peer-Based Research Mentorship Programme and Protocol Presentation Competition for and by the Medical Undergraduate Students in India - A 2 year Experience

**DOI:** 10.1101/2025.04.03.25325199

**Authors:** Shirish Rao, Devansh Lalwani, Yashika Zagade, Anveshi Nayan, Raakhi Tripathi

**Affiliations:** Graduate, Seth G.S. Medical College and K.E.M. Hospital, Mumbai, Maharashtra, India; 4th Year Medical Student, Seth G.S. Medical College and K.E.M. Hospital, Mumbai, Maharashtra, India; Intern, Seth G.S. Medical College and K.E.M. Hospital, Mumbai, Maharashtra, India; 3^rd^ Year Resident, Department of Surgery, Seth G.S. Medical College and K.E.M. Hospital, Mumbai, Maharashtra, India; Department of Pharmacology and Therapeutics, Seth G.S. Medical College and K.E.M. Hospital, Mumbai, Maharashtra, India

**Keywords:** Peer-based research mentorship, Medical undergraduate research, Research protocol development, Collaborative learning

## Abstract

**Introduction:** In response to the lack of research engagement among medical undergraduates, ASPIRE at Seth GSMC and KEM Hospital introduced an innovative mentorship program during their academic fest, Confluence. This program counters the typical challenges faced by students, such as mentorship access and practical experience, by providing structured, comprehensive training in research methodologies.

**Methodology:** In the study, four custom-developed and validated questionnaires assessed biomedical research knowledge, attitudes, skills, and post-session feedback among participants of the National Level Peer-Based Research Mentorship Programme (NLPRMP) in India. The prospective evaluation study included mentees from the 2021 and 2022 cohorts, using a rigorous selection process for team formation based on research interests and experience. The program curriculum, revised in 2022 based on feedback, encompassed key research methodology topics and concluded with a protocol presentation competition. Data analysis involved pre and post-programme questionnaires, using statistical tools to evaluate shifts in participants’ research perspectives and capabilities.

**Results:** The study, involving 64 medical students in a mentorship program, demonstrated significant improvements in various areas post-mentorship. Knowledge scores increased notably, attitudes towards medical research became more positive, motivation for engaging in medical research grew, and there were enhancements in research-related skills. Additionally, students’ perception of research as a career option and their views on integrating research into the medical curriculum improved substantially, underlining the mentorship program’s profound impact on their academic and professional development.

**Conclusion:** Exponere, the National Level Peer-Based Research Mentorship Programme at Seth GSMC and KEM Hospital significantly enhanced research engagement among medical undergraduates. It effectively improved their knowledge, attitudes, and skills in biomedical research, fostering a greater inclination towards research as a career path. This innovative approach, addressing the previously identified gaps in early research exposure and practical experience, underscores the program’s pivotal role in shaping the future of evidence-based medical practice.

## Introduction

Exposing medical students to research methodologies early in their education is a critical step in cultivating analytical thinking and guiding them towards a career in evidence-based medicine (1). This importance is underscored by findings from a systematic review by Straus et al., which indicates that medical students involved in school publications are more likely to pursue an academic career (2). The growing intricacies of modern medicine further highlight a significant issue: the lack of interest in research among medical trainees, particularly during their foundational undergraduate years (3). Numerous studies have demonstrated the positive impact of early exposure to research, showing it can effectively inspire and engage medical professionals in research activities (4)(5).

In the specific context of Indian medical education, particularly the Bachelor of Medicine, Bachelor of Surgery (MBBS) program, there’s a distinct lack of structured theoretical and practical research training for undergraduate students. Consequently, academic research often becomes a secondary, extracurricular endeavor, forcing students to seek knowledge and experience beyond their rigorous coursework independently. Research focusing on the attitudes and practices of undergraduate medical students towards research points to a widespread disinterest and lack of active involvement (6)(7)(8). Additionally, those students who initially demonstrate an interest in research frequently encounter substantial barriers such as difficulty in finding mentors, limited access to necessary resources, and a lack of practical research experience. These challenges often lead to either withdrawal from research projects or the production of substandard research work (6)(7)(8)(9).

In response to these challenge, Association for Support & Propagation of Innovation, Research & Education (A.S.P.I.R.E), student’s research council of Seth GSMC and KEM Hospital took a proactive step by launching Exponere, an innovative initiative during their academic fest, ‘Confluence’. This initiative was inspired from the already ongoing peer mentorship model at the institute and was strategically scaled-up to address and fill the gaps in research skills and knowledge among emerging medical professionals across the country.(10) It was recognized that a structured, holistic approach to research mentorship was necessary to adequately prepare and empower the next generation of medical researchers. This program was meticulously designed to provide the essential tools, guidance, and insights needed for medical students to excel in the field of research, ultimately contributing to the advancement of evidence-based medical practice.

## 2. Methods

### Study Design

The research employed a prospective evaluation study methodology, seeking to meticulously analyze the experiences and outputs of mentees who participated in and successfully completed the National Level Peer-Based Research Mentorship Programme (NLPRMP) during the span of 2021 and 2022. All the mentees who were recruited into the National Level Peer-Based Research Mentorship Programme and successfully completed it were included. Participants who dropped out of the programme midway were excluded.

### Ethical Consideration

The study was approved by the Institutional Ethics Committee (II) of Seth G.S. Medical College and K.E.M. Hospital, Mumbai, India (EC/OA-42/2021). There were no amendments or deviations in the protocol during the study period. Signed informed consent was obtained from the participants before enrolling them into the study. Participants were free to drop out or withdraw consent from the study at any point during the study period.

### Participant Selection and Team Formation

Total 39 participants were recruited from NLPRMP 2021 and 25 participants were recruited from NLPRMP 2022. Applications for mentors and mentees were received from all over India. Participants were carefully chosen through a stringent selection process involving a review of their curriculum vitae followed by a brief interview. The selection panel evaluated applicants based on criteria including, but not limited to, their educational background, past research engagements, and stated fields of interest.

To foster a rich learning environment and facilitate peer-based learning, successful applicants were then organized into diverse teams. Each team, containing four to five mentees, was structured to amalgamate individuals with converging research interests but varying levels of research experience. This ensured a mixture of expertise levels, encouraging a collaborative and enriching learning experience. Leading each team was a mentor with substantial experience in research, ranging from final-year undergraduates to postgraduates, steering the mentees through the research journey.

### Mentorship Structure and Curriculum

Throughout the duration of the program, mentors undertook the responsibility of guiding their mentees through a rich curriculum encompassing various practical and theoretical elements of basic research methodology. To aid this educational endeavor, comprehensive study material was distributed amongst the teams. While mentors had the freedom to choose their teaching content, covering core topics was essential: crafting a research question, conducting a literature search, understanding different study designs, designing a questionnaire, developing a research protocol, and understanding IEC, GCP, and ethical considerations in research.

To maintain a structured learning journey, teams were required to maintain and submit a weekly log detailing the scheduled and accomplished tasks. The program followed a dynamic yet structured four-week plan as summarized in **Table 1**. In the second year of the National Language Processing Research Mentorship Program (NLPRMP) implementation in 2022, modifications were instituted, drawing upon experiences and feedback garnered from participants during its inaugural year. Firstly, the program duration was expanded from four to five weeks, accommodating a one-week buffer to facilitate rapport and coordination development among team members. Secondly, the number of teams was strategically reduced from 13 to 5 to streamline organizational processes. Thirdly, each team was assigned a coordinator to assure the punctual attainment of weekly objectives and the execution of teaching activities by the mentor. Lastly, mentors were furnished with monetary incentives upon program completion, enhancing motivational aspects.

**Table 1.**
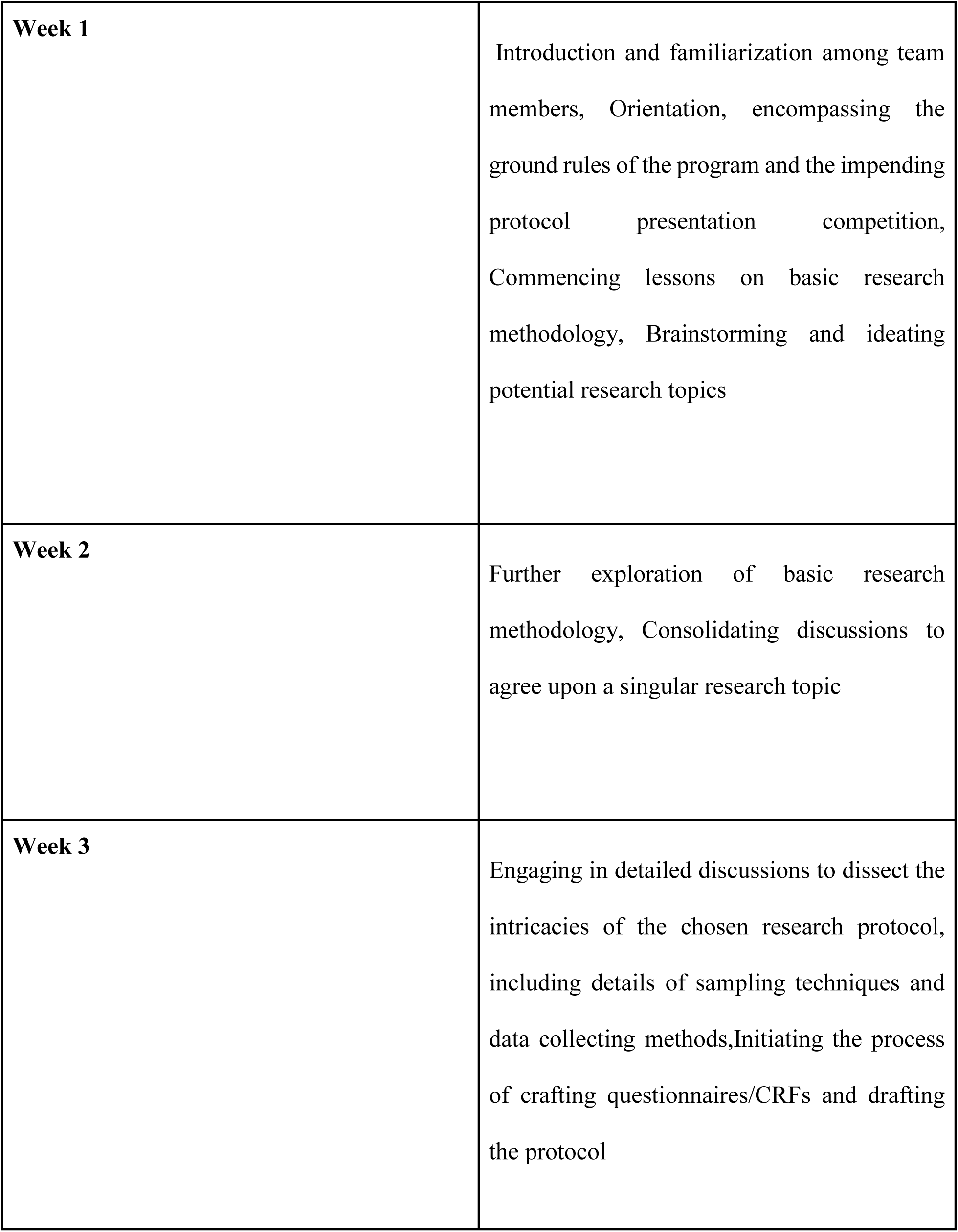
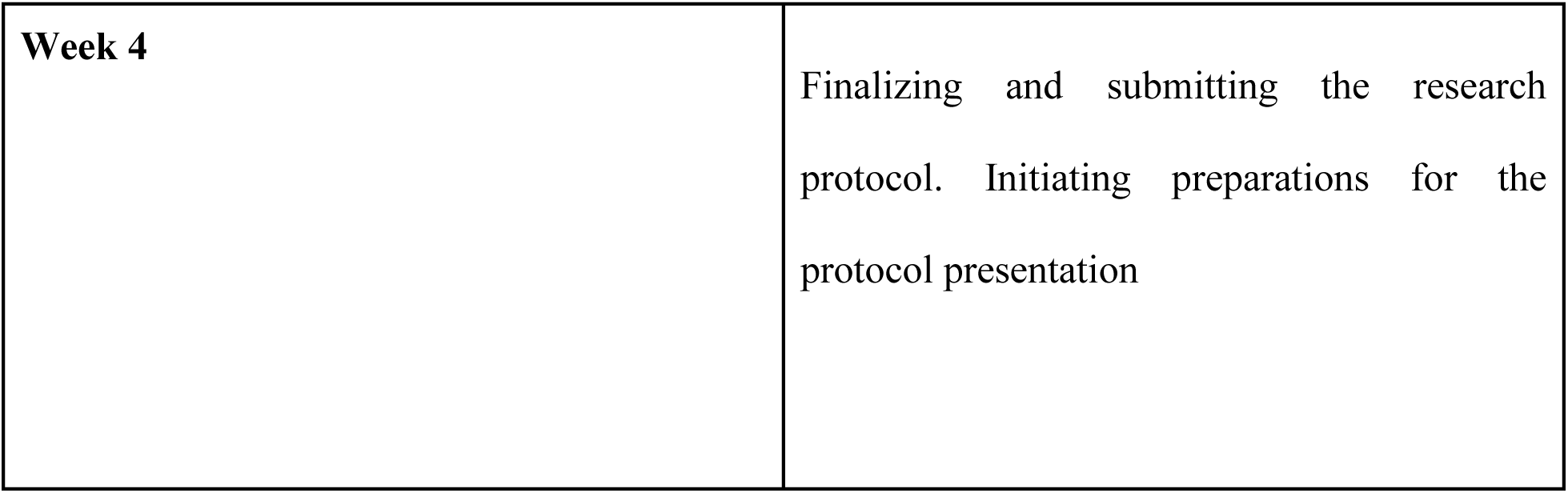
The weekly schedule followed in the course

|  |  |
| --- | --- |
| **Week 1** | Introduction and familiarization among team members, Orientation, encompassing the ground rules of the program and the impending protocol presentation competition, Commencing lessons on basic research methodology, Brainstorming and ideating potential research topics |
| **Week 2** | Further exploration of basic research methodology, Consolidating discussions to agree upon a singular research topic |
| **Week 3** | Engaging in detailed discussions to dissect the intricacies of the chosen research protocol, including details of sampling techniques and data collecting methods,Initiating the process of crafting questionnaires/CRFs and drafting the protocol |
| **Week 4** | Finalizing and submitting the research protocol. Initiating preparations for the protocol presentation |

### Protocol Presentation Competition

The culmination of the mentorship programme was a protocol presentation competition where each team presented their novel research protocol to a panel of distinguished researchers and scientists. The panel was divided into a common judging panel and subject-expert panels, providing feedback and scoring based on uniform criteria that assessed the quality and feasibility of the proposed research, including aspects such as budget and potential hurdles. This stage not only showcased the accumulation of their learning but also demonstrated collaborative learning in a competitive setting, bringing the initiative to a fruitful completion.

The jury was composed of two specialized panels: a consistent judging panel, universally applicable to all presentations, and a subject expert panel, meticulously selected for each respective subject presentation. This bifurcation aimed to ensure a holistic evaluation, marrying generalized assessment criteria with subject-specific expertise to confer nuanced and comprehensive feedback for each presentation.

After the Protocol Presentation, student teams were matched with senior mentors who were part of the specialized panel in order to guide them further in execution of their proposed projects in terms of data collection, analysis, manuscript writing and publication. This ensured continued mentorship from student mentors as well as senior experts for completion of the projects.

### Data Collection: Pre and Post Programme Questionnaires

To evaluate the programme’s efficacy and gather data on mentees’ learning progression, questionnaires were administered at two junctures: at the onset and at the end of the programme. The initial questionnaire, ‘Biomedical Research Knowledge Test’, evaluated the mentees’ existing knowledge on research methodology through a series of multiple-choice questions. The same questionnaire was administered at the programme’s end coupled with a ‘Retro-Pre Questionnaire’ which was utilized to appraise shifts in the participants’ attitudes, motivation, and perceived skill sets over the course of the programme. This comprehensive tool incorporated Likert scales, yes/no queries, and multiple-choice questions (MCQ), the latter carefully designed to encompass a broad range of potential responses to accurately capture participants’ experiences and learning trajectories. Change in knowledge was assessed by comparing the pre and post scores of the MCQ test and Change in attitudes, motivation and self perceived skill was assessed by analyzing the difference between the scores of the self reported retro-pre likert scale. All the administered questionnaires were validated by 5 subject experts who rated each item on a scale of 0-2 and the Content Validity Ratio was calculated to be 0.90. The questionnaire assessed the parameters enlisted in **Table 2**.

**Table 2:** Study Tools and Outcome variables

| **Tools** | **Description** | **Outcome variable Assessed** |
| --- | --- | --- |
| Biomedical Research<br>Knowledge Test | 30 items, MCQ type<br>Single best response,<br>Scoring Range 0-30, 1<br>point for each correct<br>answer | Change in Biomedical research<br>knowledge |
| Retro-Pre-Questionnaire for attitudes, motivation & skill development towards medical research | 45 items, Double 7-point Likert scale (Retro-Pre), Divided into 3 sections<br>Attitudes (17), Scoring Range 17-119<br>Motivations (14) Scoring Range 14 -98<br>Perceived Skills (14) 14-98 | Change in Research attitudes, motivation & skill |

### Data Analysis

Data was analyzed using Microsoft Excel 2019 and JASP (V.0.16.00) software. Visualizations were created using Microsoft Powerpoint and JASP (V.0.16.00) software. Descriptive statistics were performed to obtain frequencies, percentages, mean ± SD and median (IQR) of scores obtained from the questionnaires. Shapiro Wilk’s test was used to assess the normality of the scores. Based on the normality, paired T test or Wilcoxon’s Sign Rank test was used to assess the change in scores over the study period at a significance level of *P*<0.05.

## Results

In the study, 64 medical students aged 18-24 years, with an even gender split (32 females, 32 males), and participated in a mentorship program. Specifically, the examined cohort, aged 19 to 22 years, consisted of 8 females and 6 males. Summary of change in outcome variables and qualitative feedback from participants is provided in **Table 3** and **Panel 1** respectively.

**Table 3:**
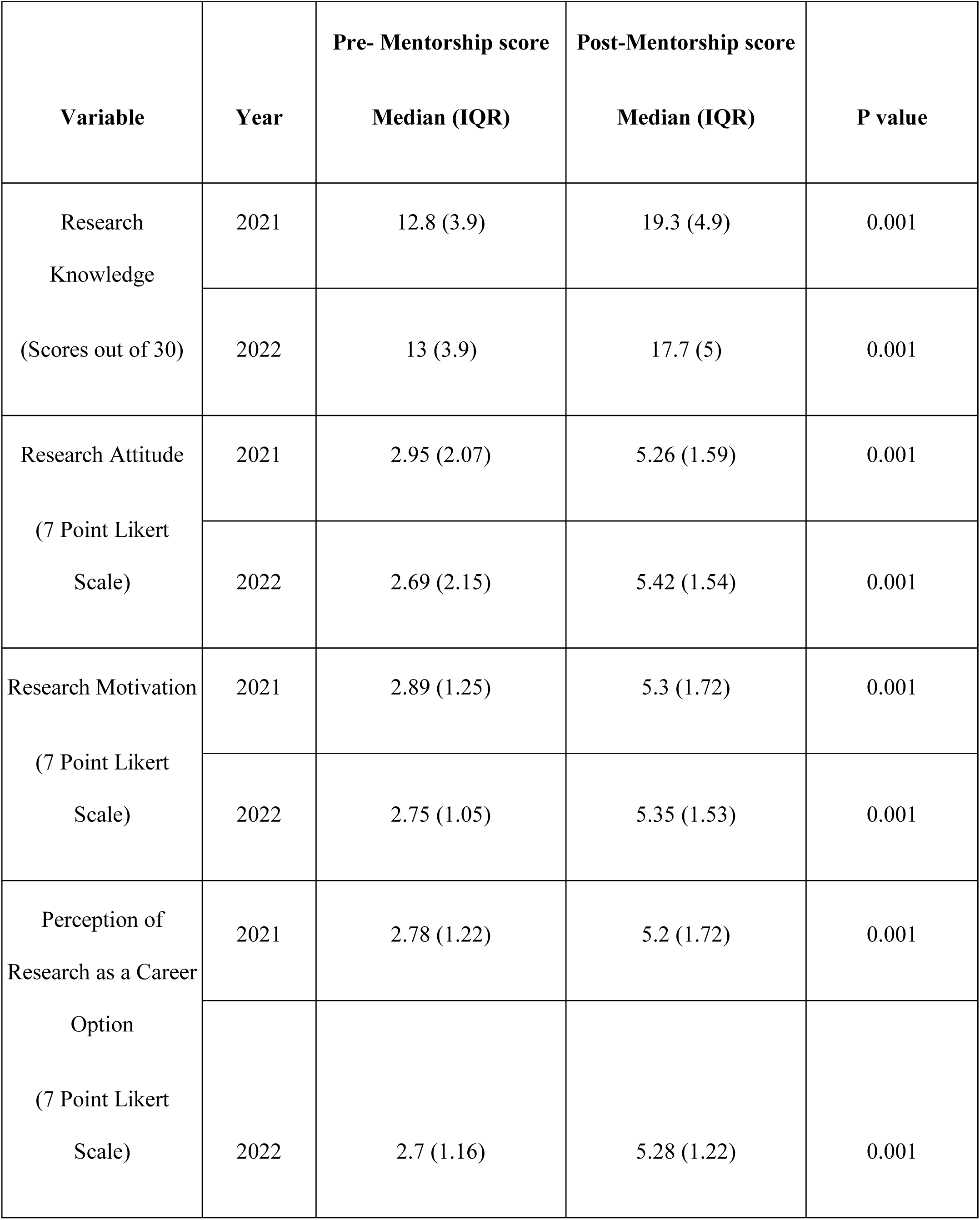
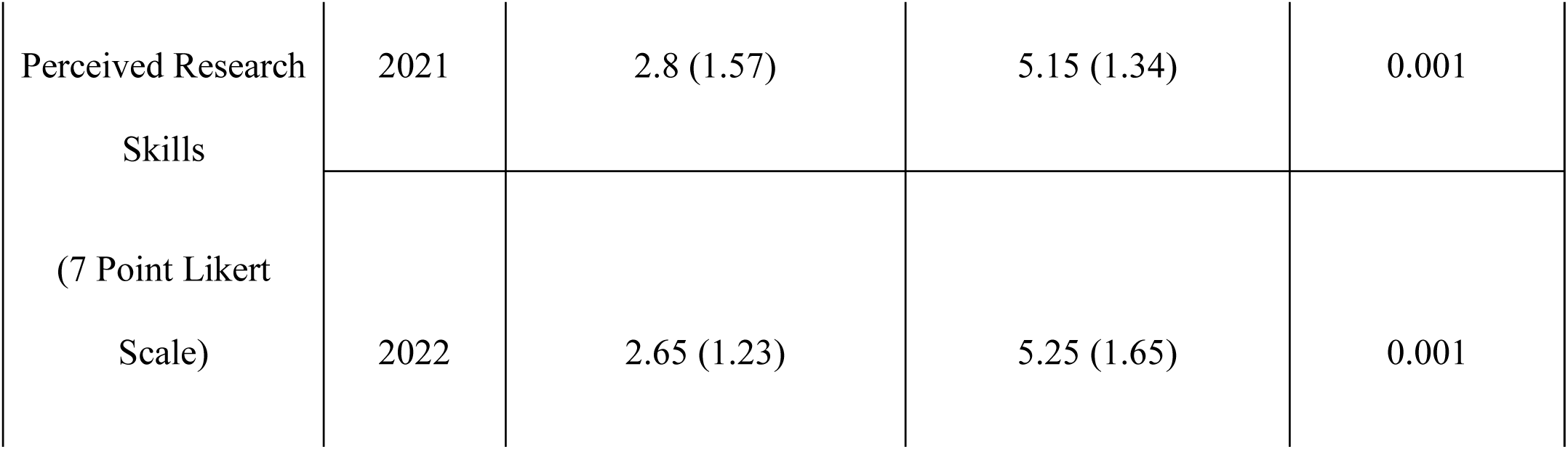
Change in scores of participants pre and post mentorship each year.

### Panel: Qualitative feedback from participants

##### Feedback from a Second-Year MBBS Student

“Joining the Exponere was a transformative experience. Initially, research seemed overwhelming, but this program broke it down into manageable and engaging components. Learning from peers who have walked the same path was incredibly relatable and effective. This experience has not only increased my research skills but also my enthusiasm for pursuing medical research.”

##### Feedback from a Third-Year MBBS Student

“Before Exponere, research felt abstract and intimidating. This program demystified it, offering practical tools and insights that made research approachable and stimulating. The peer mentorship aspect was particularly beneficial, making the learning process more collaborative and less hierarchical. I now view research as an integral part of my medical education and career.”

##### Feedback from a Final-Year MBBS Mentor

“Mentoring fellow undergraduates in Exponere was an enriching and unique experience. Seeing their growth from tentative learners to confident researchers was rewarding. The program underscores the importance of peer-led learning in research, and I was thrilled to contribute to this. It’s gratifying to see how we, as students, can facilitate each other’s academic and professional development.”

##### Feedback from a Final-Year MBBS Mentor

“Serving as a mentor in the Exponere was both a challenge and a privilege. Guiding my peers through their research journey and witnessing their development was a profound experience. The program’s peer-based structure promotes a supportive and collaborative learning environment, which I believe is crucial for cultivating a research mindset among medical students. It was inspiring to be part of a movement that empowers students to become proactive researchers early in their careers.”

#### Knowledge

In 2021, a significant improvement in knowledge scores was observed (p<0.001), with mean scores rising from 12.8 (SD 3.9) to 19.3 (SD 4.9), and median scores increasing from 4.214 to 4.86. Similarly, in 2022, the mean knowledge scores improved significantly from 13 (SD 3.9) to 17.7 (SD 5).

#### Attitude toward Medical Research

The analysis revealed a significant shift in students’ attitudes toward medical research. Pre-mentorship, the mean attitude score was 2.95 (SD=2.07, IQR=3.0) in 2021 and 2.69 (SD=2.15, IQR=4.0) in 2022. Post-mentorship, there was a notable increase to 5.26 (SD=1.59, IQR=3.0) in 2021 and 5.42 (SD=1.54, IQR=2.0) in 2022. The paired T-test showed highly significant changes (p < 0.0001), indicating a substantial positive impact of the mentorship on students’ attitudes towards medical research.

#### Motivation for Engaging in Medical Research

Students’ motivation for engaging in medical research also improved significantly after the mentorship program. The pre-mentorship mean scores were 2.89 in 2021 and 2.75 in 2022, which increased to 5.30 and 5.35 respectively post-mentorship. The changes were statistically significant (p < 0.0001), reflecting the effectiveness of the mentorship in enhancing students’ motivation towards research activities.

#### Perception of Research as a Career Option

The perception of research as a viable career option showed a marked improvement. In 2021, the pre-mentorship mean was 2.78, which increased to 5.20 post-mentorship. Similarly, in 2022, the increase was from 2.70 pre-mentorship to 5.28 post-mentorship. These changes were statistically significant (p < 0.0001), highlighting the mentorship’s influence in shaping students’ career perspectives toward research.

#### Views on Integrating Research into the Medical Curriculum

Finally, the mentorship significantly altered students’ views on integrating research into the medical curriculum. The mean scores rose from 2.83 (2021) and 2.68 (2022) pre-mentorship to 5.22 and 5.38 post-mentorship. The statistical significance of these changes (p < 0.0001) suggests a strong shift in opinion favoring the inclusion of research in medical education.

Overall, these results indicate that the mentorship program had a profound and statistically significant impact on medical students’ attitudes, motivations, skills, career perceptions, and views on the importance of research in medical education.

#### Skills Development through Medical Research

The mentorship program significantly enhanced students’ skills related to medical research. The pre-mentorship scores averaged at 2.80 (2021) and 2.65 (2022), which increased to 5.15 and 5.25 post-mentorship. The statistical analysis indicated a highly significant improvement (p < 0.0001), demonstrating the program’s role in skill development.

## Discussion

The mentorship program’s impact on medical students’ research capabilities is profound and multifaceted. The significant increase in knowledge scores indicates not only an enhancement of factual understanding but also suggests an improved ability to assimilate and apply research concepts. The shift in attitudes toward medical research is equally noteworthy. Initially moderate, these attitudes have undergone a remarkable transformation, suggesting that the mentorship program has effectively fostered a deeper appreciation for research. This is critical in an educational environment where fostering a research mindset is key to advancing medical science. Furthermore, the rise in motivation post-mentorship suggests that the program has successfully ignited a passion for research among students, which is crucial for sustaining long-term engagement in research activities.

The positive outcomes of this study align with and contribute to the growing body of research on mentorship in medical education. The work of Mulkalwar et al. (2022) (10), Rahman et al. (2021) (11) and Unnikrishnan et al. (12) (2022) highlights the importance of embedding research into medical training, a concept mirrored in our findings of improved research attitudes and skills. Moreover, innovative mentorship models, such as those explored by Maroo (2022) (13) and Bard et al. (2023) (14), reflect a paradigm shift towards more dynamic and effective research education methodologies. These parallels emphasize the crucial role of mentorship in not only enhancing academic knowledge but also in cultivating a research-oriented culture within medical education.

The study’s implications extend beyond its immediate findings. The structured nature of the mentorship program, coupled with the positive feedback from participants, positions it as a potentially scalable model that could revolutionize medical research education. It suggests a need for a systemic reevaluation of how research is integrated into medical curricula, potentially influencing future educational policies. The study also opens avenues for further research into the long-term impacts of such mentorship programs, particularly in terms of career trajectories and research outputs of medical graduates. This study serves as a catalyst for a broader conversation about the role of research in medical education, especially in regions like India, where there is a growing need for innovation in medical training. Further, the scalability of such a program is highlighted by two National level student organizations and two medical colleges from Southern India adopting its model and conducting it as a part of their own annual undergraduate medical conferences.

## Limitations

The evaluation of the “Exponere” program, while promising, must acknowledge certain constraints. Its annual occurrence and the limited number of iterations to date may not fully capture the program’s long-term efficacy. Additionally, the potential for selection bias, favoring individuals already inclined towards research, and the inherent limitations of self-reported data in accurately gauging shifts in attitudes and knowledge, should be considered when interpreting the findings.

## Conclusion

Exponere, the National Level Peer-Based Research Mentorship Programme at Seth GSMC and KEM Hospital significantly enhanced research engagement among medical undergraduates. It effectively improved their knowledge, attitudes, and skills in biomedical research, fostering a greater inclination towards research as a career path. Novelty of this initiative is that mentorship programmes are more common in research for post graduates and this particular program was conducted in undergraduate students. This innovative approach, addressing the previously identified gaps in early research exposure and practical experience, underscores the program’s pivotal role in shaping the future of evidence-based medical practice.

## Data Availability

All data produced in the present study are available upon reasonable request to the authors

